# Coast-to-coast spread of SARS-CoV-2 in the United States revealed by genomic epidemiology

**DOI:** 10.1101/2020.03.25.20043828

**Authors:** Joseph R. Fauver, Mary E. Petrone, Emma B. Hodcroft, Kayoko Shioda, Hanna Y. Ehrlich, Alexander G. Watts, Chantal B.F. Vogels, Anderson F. Brito, Tara Alpert, Anthony Muyombwe, Jafar Razeq, Randy Downing, Nagarjuna R. Cheemarla, Anne L. Wyllie, Chaney C. Kalinich, Isabel Ott, Joshua Quick, Nicholas J. Loman, Karla M. Neugebauer, Alexander L. Greninger, Keith R. Jerome, Pavitra Roychoudhury, Hong Xie, Lasata Shrestha, Meei-Li Huang, Virginia E. Pitzer, Akiko Iwasaki, Saad B. Omer, Kamran Khan, Isaac I. Bogoch, Richard A. Martinello, Ellen F. Foxman, Marie L. Landry, Richard A. Neher, Albert I. Ko, Nathan D. Grubaugh

## Abstract

Since its emergence and detection in Wuhan, China in late 2019, the novel coronavirus SARS-CoV-2 has spread to nearly every country around the world, resulting in hundreds of thousands of infections to date. The virus was first detected in the Pacific Northwest region of the United States in January, 2020, with subsequent COVID-19 outbreaks detected in all 50 states by early March. To uncover the sources of SARS-CoV-2 introductions and patterns of spread within the U.S., we sequenced nine viral genomes from early reported COVID-19 patients in Connecticut. Our phylogenetic analysis places the majority of these genomes with viruses sequenced from Washington state. By coupling our genomic data with domestic and international travel patterns, we show that early SARS-CoV-2 transmission in Connecticut was likely driven by domestic introductions. Moreover, the risk of domestic importation to Connecticut exceeded that of international importation by mid-March regardless of our estimated impacts of federal travel restrictions. This study provides evidence for widespread, sustained transmission of SARS-CoV-2 within the U.S. and highlights the critical need for local surveillance.

## Introduction

A novel coronavirus, known as SARS-CoV-2, was identified as the cause of an outbreak of pneumonia in Wuhan, China, in December 2019 (Gorbalenya et al., 2020; Wu et al., 2020; Zhou et al., 2020). Travel-associated cases of the disease COVID-19 were reported outside of China as early as January 13, 2020 and the virus has subsequently spread to nearly all nations (World Health Organization, 2020a, 2020b). The first detection of SARS-CoV-2 in the United States was a travel-associated case from Washington state on January 19, 2020 (Centers for Disease Control and Prevention, 2020a). The majority of the early COVID-19 cases in the U.S. were either i) associated with travel to a “high-risk” country or ii) close contacts of previously identified cases, per the testing criteria adopted by the Centers for Disease Control and Prevention (CDC) (Centers for Disease Control and Prevention, 2020b). In response to the risk of more travel-associated cases, the U.S. placed travel restrictions on multiple countries with SARS-CoV-2 transmission, including China on January 31, Iran on February 29, and Europe on March 11 (Taylor, 2020). However, community transmission of SARS-CoV-2 was detected in the U.S. in late February when a California resident contracted the virus despite meeting neither testing criteria (Moon et al., 2020).

From March 1 to 19, 2020, the number of reported COVID-19 cases in the U.S. rapidly increased from 74 to 13,677, and the virus was detected in all 50 U.S. states (Dong et al., 2020). It was recently estimated that the true number of COVID-19 cases in the U.S. is likely in the tens of thousands (Perkins et al., 2020), suggesting substantial undetected infections and spread within the country. We hypothesized that, with the growing number of COVID-19 cases in the U.S. and the large volume of domestic travel, new U.S. outbreaks are now more likely to result from interstate rather than international spread.

Due to its proximity to several high-volume airports, southern Connecticut is a suitable location in which to test this hypothesis. By sequencing SARS-CoV-2 from local cases and comparing their relatedness to virus genome sequences from other locations, we used ‘genomic epidemiology’ (Grubaugh et al., 2019a) to identify the likely sources of SARS-CoV-2 in Connecticut. We supplemented our viral genomic analysis with airline travel data from major airports in southern New England to estimate the risk of domestic and international importation therein. Our data suggest that the risk of domestic importation of SARS-CoV-2 into this region far outweighs that of international introductions regardless of federal travel restrictions, and find evidence for coast-to-coast U.S. SARS-CoV-2 spread.

## Results

### Phylogenetic clustering of Connecticut SARS-CoV-2 genomes demonstrates interstate spread

To delineate the roles of domestic and international virus spread in the emergence of new U.S. COVID-19 outbreaks, we sequenced SARS-CoV-2 viruses collected from cases identified in Connecticut. Our phylogenetic analyses showed that the outbreak in Connecticut was caused by multiple virus introductions and that most of these viruses were related to those sequenced from other U.S. states rather than international locations (**Figure 1**).

**Figure 1.**
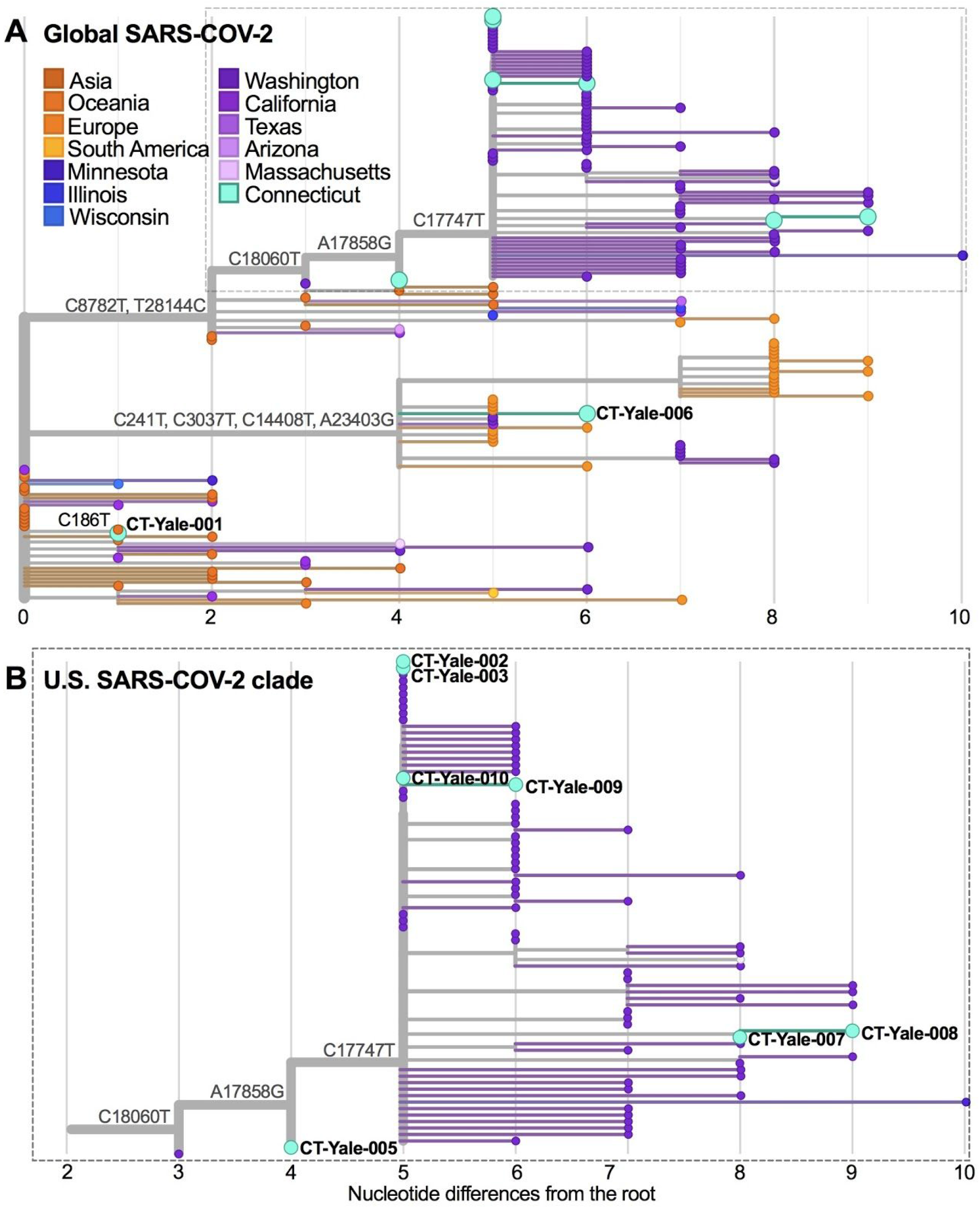
The COVID-19 outbreak in Connecticut is phylogenetically linked to SARS-CoV-2 from Washington. (**A**) We constructed a maximum-likelihood tree using 168 global SARS-CoV-2 protein coding sequences, including 9 sequences from COVID-19 patients identified in Connecticut from March 6-14, 2020. The total number of nucleotide differences from the root of the tree quantifies evolution since the putative SARS-CoV-2 ancestor. We included clade-defining nucleotide substitutions to directly show the evidence supporting phylogenetic clustering. (**B**) We enlarged the U.S. clade consisting primarily of SARS-CoV-2 sequences from Washington state and Connecticut. The MinION sequencing statistics are enumerated in **Data S1**, and the SARS-CoV-2 sequences used and author acknowledgements can be found in **Data S2**. The genomic data can be visualized and interacted with at: https://nextstrain.org/community/grubaughlab/CT-SARS-CoV-2.

We sequenced SARS-CoV-2 genomes from nine of the first COVID-19 cases reported in Connecticut (CT), with sample collection dating from March 6-14, 2020 (**Data S1**). These individuals are residents of eight different cities in Connecticut. According to the Connecticut State Department of Public Health, none of the cases were associated with international travel. Using our amplicon sequencing approach, ‘PrimalSeq’ (Grubaugh et al., 2019b; Quick et al., 2017), with the portable Oxford Nanopore Technologies (ONT) MinION platform, we generated the first SARS-CoV-2 genome approximately 14 hours after receiving the sample (CT-Yale-006), demonstrating our ability to perform near real-time clinical sequencing and bioinformatics. Our complete workflow included RNA extraction, PCR testing, validation of PCR results, library preparation, sequencing, and live base calling and read mapping. We shared the genomes of these viruses publicly as we generated them (GISAID EPI_ISL_416416-416424). We combined our genomes with other publicly available sequences for a final dataset of 168 SARS-CoV-2 genomes (**Figure 1, Data S2**). The dataset can be visualized on our ‘community’ Nextstrain page (https://nextstrain.org/community/grubaughlab/CT-SARS-CoV-2).

We built phylogenetic trees using a maximum likelihood reconstruction approach, and we used shared nucleotide substitutions to assess clade support (**Figure 1**). Our first nine SARS-CoV-2 genomes clustered into three distinct phylogenetic clades, indicating multiple independent virus introductions into Connecticut. Two of the genomes, CT-Yale-001 and CT-Yale-006, clustered primarily with viruses from China and Europe, respectively (**Figure 1A**). However, neither of the corresponding COVID-19 cases were travel-associated, which indicates that these patients were part of domestic transmission chains that stemmed from recent international virus introductions. The other seven SARS-CoV-2 genomes clustered with a large, primarily U.S. clade, within which the majority of genomes were sequenced from cases in Washington state (**Figure 1B**). Due to a paucity of SARS-CoV-2 genomes from other regions within the U.S., we could not determine the exact domestic origin of these viruses in Connecticut. We also cannot yet determine whether the higher number of substitutions observed in CT-Yale-007 and CT-Yale-008 (**Figure 1B**) compared to the other Connecticut virus genomes within this clade was the result of multiple introductions or of significant undersampling. Importantly, however, our data indicate that by early to mid March there had already been interstate spread during the early COVID-19 epidemic in the U.S.

### Travel and epidemiological patterns reveal significant domestic importation risk

Our phylogenetic analysis shows that the COVID-19 outbreak in Connecticut was driven, in part, by domestic virus introductions. To compare the roles of interstate and international SARS-CoV-2 spread in the U.S., we used airline travel data and the epidemiological dynamics in regions where travel routes originated to evaluate importation risk. We found that, due to the large volume of daily domestic air passengers, the dominant importation risk into the Connecticut region switched from international to domestic by early to mid March (**Figure 2**).

**Figure 2.**
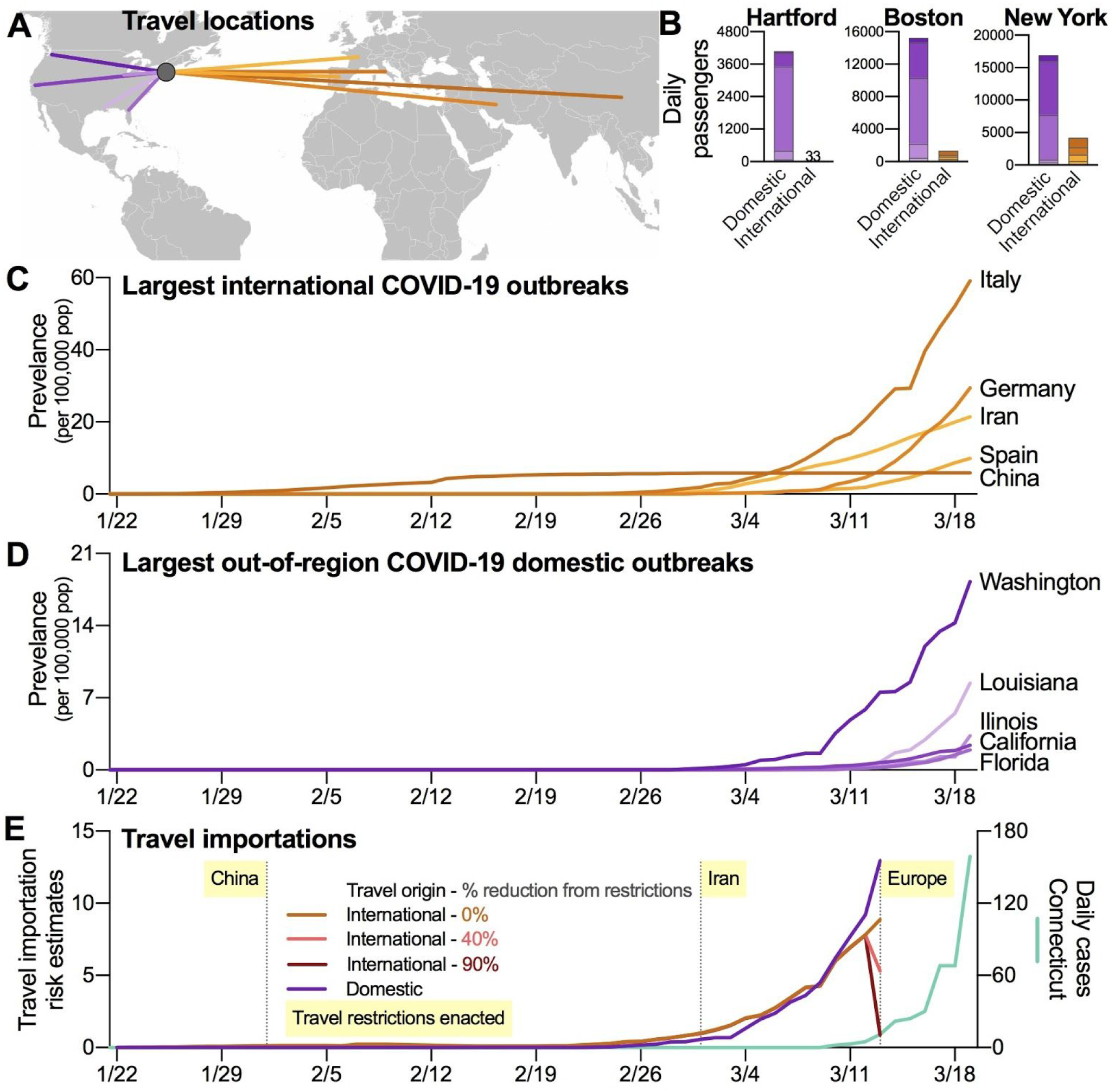
Domestic outbreaks and travel are a rising source of SARS-CoV-2 importations. (**A**) To compare the relative risks of SARS-CoV-2 importations from domestic and international sources, we selected five international (China, Italy, Iran, Spain, and Germany) and out-of-region U.S. states (Washington, California, Florida, Illinois, and Louisiana) with the highest number of reported COVID-19 cases as of March 19, 2020. (**B**) We selected three international airports in the region that are commonly used by Connecticut residents: Hartford (BDL), Boston (BOS), and New York (JFK). We used data from January to March, 2019, to estimate relative differences in daily air passenger volumes from the selected origins to the airport destinations. These daily estimates were then combined by either international or domestic travel. (**C-D**) The cumulative number of daily COVID-19 cases were divided by 100,000 population to calculate normalized disease prevalence for each location. (**E**) We calculated importation risk by modelling the number of daily prevalent COVID-19 cases in each potential importation source and then estimating the number of infected travelers using the daily air travel volume from each location. Data, criteria, and analyses used to create this figure can be found in **Data S3**.

We first estimated daily passenger volumes arriving in the region from the five countries (China, Italy, Iran, Spain, and Germany) and out-of-region states (Washington, California, Florida, Illinois, and Louisiana) that have reported the most COVID-19 cases to date (**Figure 2A-D**). To this end, we collected passenger volumes arriving in three major airports in southern New England: Bradley International Airport (BDL; Hartford, Connecticut), General Edward Lawrence Logan International Airport (BOS; Boston, Massachusetts), and John F. Kennedy International Airport (JFK; New York, New York; **Figure 2B**). As travel data for 2020 are not yet available, we calculated the total passenger volume from each origin and destination pair between January and March, 2019, and estimated the number of daily passengers. We found that the daily domestic passenger volumes were ∼100 times greater than international in Hartford, ∼10 times greater in Boston, and ∼4 times greater in New York in our dataset (**Figure 2B**).

By combining daily passenger volumes (**Figure 2B**) with COVID-19 prevalence at the travel route origin (**Figure 2C-D**) and accounting for differences in reporting rates, we found that both domestic and international SARS-CoV-2 importation started to increase dramatically at the beginning of March, 2020 (**Figure 2E**). Without accounting for the effects of international travel restrictions, our estimated domestic importation risk from the selected five U.S. states surpassed international importation risk by March 10. Using previous assumptions around travel restrictions (Chinazzi et al., 2020), we also modeled two scenarios where federal travel restrictions reduced passenger volume by 40% and by 90% from the restricted countries (**Figure 2E**). Due to the overall low prevalence of COVID-19 in China, we did not find any significant effects of travel restrictions from China that were enacted on February 1st (**Data S3**). Also, we did not find significant changes to the importation risk following travel restrictions from Iran on March 1, likely due to the relatively small number of passengers arriving from that country (**Data S3**). While we did find a dramatic decrease in international importation risk following the restrictions on travel from Europe (March 13), this decrease occurred after our estimates of domestic travel importation risk had already surpassed that of international importation (**Figure 3E**). The dramatic rises in both domestic and international importation risk preceded the state-wide COVID-19 outbreak in Connecticut (**Figure 3E**), and the recent increase in risk of domestic importation may give rise to new outbreaks in the region.

## Discussion

The combined results of our genomic epidemiology and travel pattern analyses suggest that domestic spread recently became a significant source of new SARS-CoV-2 infections in the U.S. We find strong evidence that outbreaks on the East Coast (Connecticut) are linked to outbreaks on the West Coast (Washington), demonstrating that trans-continental spread has already occured. As of March 25, there are >1000 SARS-CoV-2 genomes sequenced from around the world, including >350 from the U.S. (https://nextstrain.org/ncov); however most of the latter were obtained from a small number of U.S. states. Therefore, we cannot yet determine the exact origins of the viral introductions into Connecticut. Recent domestic travel history of the nine reported cases was not available, but it is unlikely that all of the infections originated in Washington state. Furthermore, due to low genetic diversity between these early sequences from Connecticut and Washington, we cannot yet determine how often the virus may be spreading between the U.S. coasts or whether an introduction from a common source is responsible for phylogenetic grouping. As testing capacity increases and more viral genome sequences become available from new locations, more granular reconstructions of virus spread throughout the U.S. will be possible (Grubaugh et al., 2019a). Specifically, elucidating the phylogenetic relationship of viral genomes collected in Connecticut to those collected in neighboring states, especially those with a high burden of disease like New York, will improve our understanding of critical interstate dynamics.

Our estimates of domestic importation risk are likely conservative despite some important limitations in our air travel analysis. Because we do not have access to current airline data, we could not exactly quantify the impact of government restrictions on international travel. In addition, even without explicit government restrictions, general social distancing and work-from-home guidelines are reducing all airline travel. We did not account for these decreases in either our international or domestic travel data. While such variations may lower our domestic risk estimates, we also did not account for the large volumes of regional automobile and rail travel, especially along the corridor that connects Massachusetts, New York, New Jersey, Pennsylvania, and Washington D.C. to Connecticut. As such, the interconnectedness and large volumes of daily travel within the U.S. indicate that domestic spread of SARS-CoV-2 has become and will likely continue to be the primary source of new infections.

We argue that, though simplistic, our model demonstrates the urgent need to focus control efforts in the U.S. on preventing further domestic virus spread. In China, local outbreak dynamics were highly correlated with travel between Wuhan and the outbreak dynamics therein during the early months of the epidemic (Kraemer et al., 2020). Similarly, if interstate introductions are not curtailed in the U.S. with improved surveillance measures, more robust diagnostic capabilities, and proper clinical care, quelling local transmission within states will be a Sisyphean task. We therefore propose that a unified effort to detect and prevent new COVID-19 cases will be essential for mitigating the risk of future domestic outbreaks. This effort must ensure that states have sufficient personal protective equipment, sample collection materials, and testing reagents, as these supplies enable effective surveillance. Finally, state- and local-level policymakers must recognize that the health and well-being of their constituents are contingent on that of the nation. If spread between states is now common, as our results indicate, the U.S. will struggle to control COVID-19 in the absence of a unified surveillance strategy.

## Methods

### Lead Contact and Materials Availability

Further information and requests for data, resources, and reagents should be directed to and will be fulfilled by the Lead Contact, Nathan D. Grubaugh. This study did not generate new unique reagents, but raw data and code generated as part of this research can be found in the **Supplemental Files**, as well as on public resources as specified in the **Data and Code Availability** section below.

### Ethics Statement

Residual de-identified nasopharyngeal samples testing positive for SARS-CoV-2 by reverse-transcriptase quantitative (RT-q)PCR were obtained from the Yale-New Haven Hospital Clinical Virology Laboratory or the Connecticut State Department of Public Health. In accordance with the guidelines of the Yale Human Investigations Committee and the Connecticut State Department of Public Health, this work with de-identified samples is considered non-human subjects research. All samples were de-identified before receipt by the study investigators.

### Sample collection, processing, and sequencing

Samples for this study were collected during an early testing phase by the Connecticut State Department of Public Health or the Yale Clinical Virology Laboratory at the Yale School of Medicine. Nasopharyngeal swabs were collected from patients presenting with symptoms of SARS-CoV-2 infection at multiple medical centers in Connecticut. These patients are all Connecticut residents, but we do not have access to location data associated with each of these early SARS-CoV-2 genomes to avoid patient identification. Swabs were placed in virus transport media (BD Universal Viral Transport Medium) immediately upon collection. Samples (200 µL) were subjected to total nucleic acid extraction using the NUCLISENS easyMAG platform (BioMérieux, France) at the Yale Clinical Virology Laboratory. The recommended CDC RT-qPCR assay was used to test for the presence of SARS-CoV-2 RNA (Centers for Disease Control and Prevention, 2020c). A total of 10 samples from 10 different individuals with cycle-threshold (CT) values less than 35 were selected to move forward with next generation sequencing (NGS). Of these, we were successfully able to generate sequencing libraries from nine samples.

SARS-CoV-2 positive samples were processed for NGS using a highly multiplexed PCR amplicon approach for sequencing on the Oxford Nanopore Technologies (ONT; Oxford, United Kingdom) MinION using the V1 primer pools (Quick et al., 2017). Sequencing libraries were barcoded and multiplexed using the Ligation Sequencing Kit and Native Barcoding Expansion pack (ONT) following the ARTIC Network’s library preparation protocol (Quick, 2020) with the following minor modifications: cDNA was generated with SuperScriptIV VILO Master Mix (ThermoFisher Scientific, Waltham, MA, USA), a total of 20 ng of each sample was used as input into end repair, end repair incubation time was increased to 25 minutes followed by a 1:1 bead-based clean up, and Blunt/TA ligase (New England Biolabs, Ipswich, MA, USA) was used to ligate barcodes to each sample. cDNA synthesis and amplicon generation was performed concurrently for each sample. Samples were processed by CT value to reduce the likelihood of contamination from high titer samples to low titer samples. Barcoding, adaptor ligation, and sequencing was performed on samples with CT values between 25-35 (low titer group) prior to samples with CT values below 25 (high titer group) (**Data S1**). Two samples, Yale-006 and Yale-007, were diluted 1:100 in nuclease-free water prior to cDNA synthesis. A no template control was created at the cDNA synthesis step and amplicon generation step to detect cross-contamination between samples. Controls were barcoded and sequenced with both the high and low titer sample groups.

A total of 24 ng of the low titer group was loaded onto a MinION R9.4.1 flow cell and sequenced for a total of 5.5 hours and generated 2.1 million reads. The flow cell was nuclease treated, flushed, and primed prior to loading 25 ng of the high titer group library. These samples were sequenced for a total of 9 hours and generated 1.4 million reads (**Data S1**). The RAMPART software from the ARTIC Network was used to monitor the sequencing run to estimate the depth of coverage across the genome for each barcoded sample in both runs https://github.com/artic-network/rampart). Following completion of the sequencing runs, .fast5 files were basecalled with Guppy (v3.5.1, ONT) using the high accuracy module. Basecalling was performed on a single GPU node on the Yale HPC. Consensus genomes were generated for input into phylogenetic analysis according to the ARTIC Network bioinformatic pipeline ([Artic Network](https://artic.network/ncov-2019/ncov2019-bioinformatics-sop.html)). Variants in the consensus genomes were called using nanopolish per the bioinformatic pipeline (Loman et al., 2015). Amplicons that were not sequenced to depth of 20x were not included in the final consensus genome, and these positions are represented by stretches of NNN’s (**Data S1**).

### Phylogenetic analysis

To investigate the origin and diversity of SARS-CoV-2 in Connecticut, we compiled a dataset of our nine genomes with another 159 representative sample of SARS-CoV-2 genomes that were available from GenBank (https://www.ncbi.nlm.nih.gov/genbank/sars-cov-2-seqs/) and GISAID (https://www.gisaid.org/). See **Data S2** for a list of sequences and acknowledgements to the originating and submitting labs. No data that was only released on GISAID was used without consent from the authors (see **Acknowledgements**). We aligned consensus genomes using the augur toolkit version 6.4.2 (Hadfield et al., 2018). Specifically, we aligned sequences using mafft (Katoh et al., 2002), masked sites at the 5’ and 3’ ends of the alignment as well as a small number of sites that likely vary due to assembly artefacts (see https://github.com/nextstrain/ncov), and reconstructed a phylogeny using IQ-Tree (Nguyen et al., 2015). These trees are further processed using augur and treetime to add ancestral reconstructions (Sagulenko et al., 2018). Sequences in this sample differ from the root by 10 or fewer mutations. Bootstrap values are not a meaningful measure of branch support in this case. Instead, we directly show mutations supporting the major splits in the tree. The mutations defining these clades are compatible with the tree topology and are not homoplastic. The probability that all clade defining mutations arose multiple times independently in a manner compatible with the tree topology is vanishingly small. The data can be visualized at: https://nextstrain.org/community/grubaughlab/CT-SARS-CoV-2.

### International and U.S. COVID-19 cases

Daily COVID-19 cases from international locations were obtained from the European Centre for Disease Prevention and Control via Our World in Data (https://ourworldindata.org/coronavirus-source-data). International data were accessed on March 19, 2020. Daily COVID-19 cases from Connecticut and other U.S. locations (Washington, California, Florida, Illinois, and Louisiana) were obtained from the repository (https://github.com/CSSEGISandData/COVID-19) hosted by the Center for Systems Science and Engineering (CSSE) at Johns Hopkins University (Dong et al., 2020). These represent the international and out-of-region domestic (i.e. excluding New York, Massachusetts, and New Jersey) locations with the most reported COVID-19 cases.

### Air passenger volumes

To investigate the domestic and international spread of SARS-CoV-2, we obtained air passenger volumes from the International Air Transport Association (IATA; https://www.iata.org/pages/default.aspx). IATA data consists of global ticket sales, which account for true origins and final destinations, and represents 90% of all commercial flights. We obtained the monthly number of number of passengers traveling by air from five international (China, Italy, Iran, Spain, and Germany) and five U.S. locations (Washington, California, Florida, Illinois, and Louisiana) to airports that are commonly used by Connecticutians: Bradley International Airport (BDL, Hartford, Connecticut; ranked 53rd in U.S. in yearly passenger volume; https://www.faa.gov/airports/planning_capacity/passenger_allcargo_stats/passenger/), General Edward Lawrence Logan International Airport (BOS, Boston, Massachusetts; ranked 16th), and John F. Kennedy International Airport (JFK, New York, New York; ranked 6th). Air passenger data from 2020 is not currently available; thus, we used data from January to March 2019 to represent general trends in passenger volumes, as done previously (Bogoch et al., 2020). We took the average of the 3-month passenger volumes to estimate the daily number of travelers entering each airport from the specified origin. To account for passenger reductions following U.S. government alerts and restrictions (Taylor, 2020), we modeled two scenarios: a 40% reduction in passenger volume and a 90% reduction in passenger volume. These thresholds were determined based on previously reported estimates and assumptions around travel restrictions (Chinazzi et al., 2020).

### Travel importation risk estimates

We estimated the true number of incident cases per day by adjusting the number of reported incident cases to reflect the ascertainment period and reporting rate using

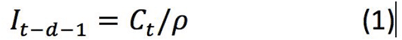

where *C*_*t*_ is the number of reported incident cases of COVID-19 on day *t, d* is the number of days from symptom onset to testing, and ρ is the reporting rate.

We assumed a constant ascertainment period of *d*=5 days between symptom onset and testing (Ferguson et al., 2020). Because of the evidence of pre-symptomatic transmission (Tindale et al., 2020), we also assumed that cases become infectious one day before symptom onset. To account for substantial uncertainty around reporting rates, we assigned different reporting rates to individual locations based on the testing criteria enacted in that location (Niehus et al., 2020). For each country and state, we first extracted testing criteria from the department or ministry of health website. We assumed that countries or states with similar testing criteria policies captured similar proportions of true infections. Using the respective testing criteria, we categorized countries or states as having narrow, moderate, or broad testing levels. We then assigned reporting rates to each testing level by using the mean and 95% confidence interval of the reporting rate estimated by Nishiura et al. (Nishiura et al., 2020): 0.092 (95% CI= 0.05–0.20). The reporting rate for the broadest testing level, ρ=0.20, also corresponded to the reporting rate in Mainland China (Chinazzi et al., 2020). We thus assigned Iran, Florida, Washington, and Illinois to a “narrow” testing level (ρ=0.05); Spain, Italy, and Louisiana to a “moderate’’ testing level (ρ=0.092); and China, Germany, and California to a “broad” testing level (ρ=0.20, **Data S2**, “testing-criteria”).

To estimate the number of prevalent infectious individuals on day *t* (*P*_*t*_), we multiplied the number of incident infections up to day *t* by the probability that an individual who became infectious on day *i* was still infectious on day *t*:

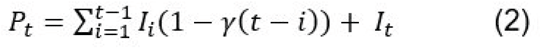

Where γ(t-i) is the cumulative distribution function of the infectious period. We modeled the infectious period as gamma distribution with mean 7 days and standard deviation 4.5 days which aligns with other modeling studies (Prem et al., 2020; Zhao et al., 2020).

We assumed that cases would not travel once they were diagnosed and therefore removed them from our estimate of infectious travelers (*T*_*t*_):

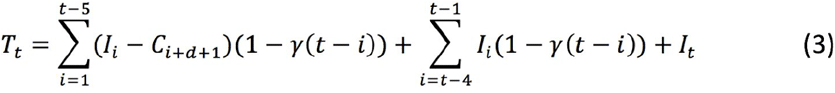

The first term of Equation 3 accounts for the assumption that some cases had been diagnosed by day *t* and thus would not travel. The second and third terms capture cases who are infectious on day *t* and have not yet been diagnosed.

We calculated daily risk of importation as a function of the population-adjusted density of infectious travelers and passenger volume:

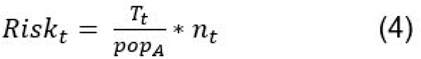

where *T*_*t*_ is the number of infectious travelers on day *t, pop*_*A*_ is the population of location *A*, and *n*_*t*_ is the number of passengers traveling from each location to southern New England on day *t*. We summed the calculated risk across the three airports (BDL, BOS, JFK) and then across domestic and international travelers to arrive at our final estimates.

### Quantification and Statistical Analysis

Statistical analyses were performed using R version 3.5.2 (R Core Team, 2017) and are described in the figure legends and in the method details.

### Data and Code Availability

The SARS-CoV-2 sequences generated here can be found using the NCBI BioProject: (PRJNA614976) and GISAID (EPI_ISL_416416-416424). Sequencing data will be made available via SRA. Data used to create the figures can be found in the supplemental files. The interactive Nextstain page to visualize the genomic data can be found at: https://nextstrain.org/community/grubaughlab/CT-SARS-CoV-2. The raw data, results, and analyses can be found at: https://github.com/grubaughlab/CT-SARS-CoV-2.

## Data Availability

https://github.com/grubaughlab/CT-SARS-CoV-2

## Acknowledgements

The authors of this study would like to acknowledge S. Cordey, I. Eckerle and L. Kaiser from Geneva University Hospital for directly sharing their genome sequence data with our team, everyone who openly shared their genomic data on GenBank and GISAID (authors listed in Data S2), D. Ferguson, R. Garner and J. Criscuolo at the Yale Clinical Virology Laboratory for their laboratory support, the staff of the Yale Center for Research Computing for technical support, S. Taylor and P. Jack for enlightening discussions, our friends, families, and the Yale community for support during this difficult time, and all of the health care workers, public health employees, and scientists for their COVID-19 response.

This research was funded by the generous support from the Yale Institute for Global Health and the Yale School of Public Health start-up package provided to NDG. CBFV is supported by NWO Rubicon 019.181EN.004. VEP is funded by NIH/NIAID R01 AI112970 and R01 AI137093. NJL is funded by a Medical Research Council fellowship as part of the CLIMB project. The ARTIC resources were funded by a Wellcome Trust Collaborative Award project number 206298/A/17/Z. JQ is funded by a UKRI Future Leaders Fellowship. KMN is funded by NIH R01 GM112766. IIB is funded by a COVID-2019 grant through the Canadian Institutes of Health Research.

## Author Contributions

## Supplemental Information

**Data S1**. MinION sequencing statistics, related to **Figure 1**.

**Data S2**. SARS-CoV-2 sequences used and acknowledgements, related to **Figure 1**.

**Data S3**. Air passenger volumes, COVID-19 prevalence, testing levels, and importation risk, related to **Figure 2**.

## References

Bogoch, I.I., Watts, A., Thomas-Bachli, A., Huber, C., Kraemer, M.U.G., and Khan, K. (2020). Potential for global spread of a novel coronavirus from China. J. Travel Med. 27.

Centers for Disease Control and Prevention (2020a). First Travel-related Case of 2019 Novel Coronavirus Detected in United States.

Centers for Disease Control and Prevention (2020b). Evaluating and Reporting Persons Under Investigation (PUI) Coronavirus Disease 2019 (COVID-19).

Centers for Disease Control and Prevention (2020c). CDC 2019-Novel Coronavirus (2019-nCoV) Real-Time RT-PCR Diagnostic Panel.

Chinazzi, M., Davis, J.T., Ajelli, M., Gioannini, C., Litvinova, M., Merler, S., Pastore Y Piontti, A., Mu, K., Rossi, L., Sun, K., et al. (2020). The effect of travel restrictions on the spread of the 2019 novel coronavirus (COVID-19) outbreak. Science.

Dong, E., Du, H., and Gardner, L. (2020). An interactive web-based dashboard to track COVID-19 in real time. Lancet Infect. Dis.

Gorbalenya, A.E., Baker, S.C., Baric, R.S., de Groot, R.J., Drosten, C., Gulyaeva, A.A., Haagmans, B.L., Lauber, C., Leontovich, A.M., Neuman, B.W., et al. (2020). Severe acute respiratory syndrome-related coronavirus: The species and its viruses – a statement of the Coronavirus Study Group.

Grubaugh, N.D., Ladner, J.T., Lemey, P., Pybus, O.G., Rambaut, A., Holmes, E.C., and Andersen, K.G. (2019a). Tracking virus outbreaks in the twenty-first century. Nat Microbiol, 10–19.

Grubaugh, N.D., Gangavarapu, K., Quick, J., Matteson, N.L., De Jesus, J.G., Main, B.J., Tan, A.L., Paul, L.M., Brackney, D.E., Grewal, S., et al. (2019b). An amplicon-based sequencing framework for accurately measuring intrahost virus diversity using PrimalSeq and iVar. Genome Biol. 20, 8.

Hadfield, J., Megill, C., Bell, S.M., Huddleston, J., Potter, B., Callender, C., Sagulenko, P., Bedford, T., and Neher, R.A. (2018). Nextstrain: real-time tracking of pathogen evolution. Bioinformatics 34, 4121–4123.

Katoh, K., Misawa, K., Kuma, K.-I., and Miyata, T. (2002). MAFFT: a novel method for rapid multiple sequence alignment based on fast Fourier transform. Nucleic Acids Res. 30, 3059–3066.

Kraemer, M.U.G., Yang, C.-H., Gutierrez, B., Wu, C.-H., Klein, B., Pigott, D.M., Open COVID-19 Data Working Group†, du Plessis, L., Faria, N.R., Li, R., et al. (2020). The effect of human mobility and control measures on the COVID-19 epidemic in China. Science.

Loman, N.J., Quick, J., and Simpson, J.T. (2015). A complete bacterial genome assembled de novo using only nanopore sequencing data. Nat. Methods 12, 733–735.

Moon, S., Yan, H., Christensen, J., and Maxouris, C. (2020). The CDC has changed its criteria for testing patients for coronavirus after the first case of unknown origin was confirmed. CNN.

Nguyen, L.-T., Schmidt, H.A., von Haeseler, A., and Minh, B.Q. (2015). IQ-TREE: a fast and effectiv 32, 268–274.

Niehus, R., De Salazar, P.M., Taylor, A., and Lipsitch, M. (2020). Quantifying bias of COVID-19 prevalence and severity estimates in Wuhan, China that depend on reported cases in international travelers (medRxiv).

Nishiura, H., Kobayashi, T., Yang, Y., Hayashi, K., Miyama, T., Kinoshita, R., Linton, N.M., Jung, S.-M., Yuan, B., Suzuki, A., et al. (2020). The Rate of Underascertainment of Novel Coronavirus (2019-nCoV) Infection: Estimation Using Japanese Passengers Data on Evacuation Flights. J. Clin. Med. Res. 9.

Perkins, A.,Cavany, S.M., Moore, S.M., Oidtman, R.J., Lerch, A., and Poterek, M. (2020). Estimating unobserved SARS-CoV-2 infections in the United States (medRxiv).

Prem, K., Liu, Y., Russell, T., Kucharski, A.J., Eggo, R.M., Davies, N., Centre for the Mathematical Modelling of Infectious Diseases COVID-19 Working Group, Jit, M., and Klepac, P. (2020). The effect of control strategies that reduce social mixing on outcomes of the COVID-19 epidemic in Wuhan, China (medRxiv).

Quick, J. (2020). nCoV-2019 sequencing protocol v1 (protocols.io.bbmuik6w).

Quick, J., Grubaugh, N.D., Pullan, S.T., Claro, I.M., Smith, A.D., Gangavarapu, K., Oliveira, G., Robles-Sikisaka, R., Rogers, T.F., Beutler, N.A., et al. (2017). Multiplex PCR method for MinION and Illumina sequencing of Zika and other virus genomes directly from clinical samples. Nat. Protoc. 12, 1261–1276.

R Core Team (2017). R: a language and environment for statistical computing. R Foundation for Statistical Computing, Vienna. https.Www.R-ProjeCt.Org.Sagulenko,

P. Puller, V., and Neher, R.A. (2018). TreeTime: Maximum-likelihood phylodynamic analysis. Virus Evol 4, vex042.

Taylor, D.B. (2020). A Timeline of the Coronavirus. The New York Times.

World Health Organization (2020a). Novel Coronavirus (2019-nCoV) Situation Report-1.

World Health Organization (2020b). Coronavirus disease 2019 (COVID-19) Situation Report-63.

Wu, F., Zhao, S., Yu, B., Chen, Y.-M., Wang, W., Song, Z.-G., Hu, Y., Tao, Z.-W., Tian, J.-H., Pei, Y.-Y., et al. (2020). A new coronavirus associated with human respiratory disease in China. Nature 579, 265–269.

Zhao, Z., Zhu, Y.-Z., Xu, J.-W., Hu, Q.-Q., Lei, Z., Rui, J., Liu, X., Wang, Y., Luo, L., Yu, S.-S., et al. (2020). A mathematical model for estimating the age-specific transmissibility of a novel coronavirus (medRxiv).

Zhou, P., Yang, X.-L., Wang, X.-G., Hu, B., Zhang, L., Zhang, W., Si, H.-R., Zhu, Y., Li, B., Huang, C.-L., et al. (2020). A pneumonia outbreak associated with a new coronavirus of probable bat origin. Nature 579, 270–273.

Ferguson NM, Laydon D, Nedjati-Gilani G, et al. (2020). Impact of non-pharmaceutical interventions (NPIs) to reduce COVID-19 mortality and healthcare demand. London: Imperial College COVID-19 Response Team, March 16, 2020. <https://www.imperial.ac.uk/media/imperial-college/medicine/sph/ide/gida-fellowships/Imperial-College-COVID19-NPI-modelling-16-03-2020.pdf>

